# Transcutaneous Vagus Nerve Stimulation in the Treatment of Long Covid-Chronic Fatigue Syndrome

**DOI:** 10.1101/2022.11.08.22281807

**Authors:** Benjamin H Natelson, Michelle Blate, Tiffany Soto

## Abstract

Many patients do not recover following Covid infection. The resulting illness is called Long Covid. Because there is no agreed upon treatment for this ailment, we decided to do an open label pilot study using non-invasive, transcutaneous stimulation of the auricular branch of the vagus nerve. Inclusion criteria required the patient to fulfill criteria for having chronic fatigue syndrome. Fourteen patients provided evaluable data. Eight of these fulfilled our requirements for treatment success. Since our criterion for a successful study was that at least a third of patients had to show a positive response to treatment, this was a successful pilot that warrants a follow up study that is appropriately sham controlled.

All authors have read and approved this manuscript for submission.

There are no competing interests and support for doing this study was via patient donations to support Dr Natelson’s research activities.

The actual data summarized in Table 1 are available upon request

---

SARS CoV-2 infection has long term consequences. One study from Hong Kong was most germane to the current Covid-19 pandemic because it provided a four year follow up of the health status of 233 patients who survived the 2003 SARS outbreak^1^: Of these, 27% fulfilled 1994 case criteria for myalgic encephalomyelitis/chronic fatigue syndrome [ME/CFS], an illness characterized by at least 6 months of fatigue producing a substantial reduction in activity plus rheumatological, infectious and neuropsychiatric symptoms^2^. The current COVID pandemic has not been different. A Scottish population study found that of 30,100 PCR positive patients not requiring hospitalization and re-evaluated at least 6 months after infection, 5% showed no improvement and an additional 42% only partial recovery^3^; these data suggest a huge world-wide medical problem. This syndrome, commonly called Long Covid, shares many of the symptoms of ME/CFS including marked fatigue, problems with attention and concentration, diffuse muscular achiness/pain and marked post-exertional malaise where even minimal effort produces a dramatic worsening of all other symptoms^4^; in fact, in our recent paper, we found that nearly half of the Long Covid patients we studied^5^ fulfilled the 1994 criteria^2^ for ME/CFS.

There is no currently agreed upon treatment for Long Covid; therefore, we were impressed by a Belgian paper reporting improved symptoms in Long Covid patients following application of non-invasive auricular vagus nerve stimulation [VNS]^6^. The study we present here was our effort to try to confirm these early findings in Long Covid. In order to reduce patient pool heterogeneity, we required patients to fulfill the 1994 case definition for CFS^2^.

## Methods

Potential subjects were contacted by phone and asked to complete a questionnaire we have used successfully for many years to identify potential research subjects with ME/CFS. Sixteen subjects who had survived Covid but remained with symptoms for at least 6 months thereafter signed our IRB-approved informed consent to participate in this study via RedCap.

Subjects did not have to stop any medication they were currently taking to be part of this trial. The major criterion for intake was that subjects had to fulfill the 1994 case definition for CFS^2^: Our way of determining this uses a 5 point Likert scale [0=none; 1 = mild=; 2 = moderate; 3 = substantial; 4 = severe; 5 = very severe] and requires at least 6 months of fatigue sufficient enough to produce at least a substantial reduction in activity in one of the following spheres: work, school, personal or social. In addition, patients had to endorse having at least a substantial problem with three of the following plus at least a moderate problem on one: sore throat; tender lymph glands; headache; myalgia; arthralgia, difficulty with attention and concentration [called brain fog] and the report that even mild exertion produces a dramatic syndromic worsening [post-exertional malaise].

On intake, subjects completed the following: visual analog scales for fatigue, brain fog and widespread pain and had to endorse at least one of these as producing a substantial problem [≥ 3 on our 0 to 5 symptom severity scale] ; SF-36 [a 36 question vehicle to determine the patient’s health related quality of life]^7^, Profile of Mood States [30 words concerning which patients provide input on a 5 point scale ranging from not at all to extremely] ^8^; the Chalder fatigue scale [11 questions with four possible answers each and allowing determination of whether subjects fulfilled criteria for Fatigue]^9^. Subjects were then mailed a Parasym device with instructions to attach the electrode to their left tragus, stimulate at whatever current was just less than being uncomfortable for 35 min a day, every day for 6 weeks. We chose the 35 min stimulation duration based on the earlier report using the same device^6^.

Subjects were called every two weeks during the 6 week trial to ask about problems related to tVNS use. At the end of the 6 week trial, subjects completed these same questionnaires as well as the Patient Global Impression of Change scale [range from +3 for marked improvement to -3 for markedly worse with 0 no change].

### *A priori* criteria for primary outcome

Improvement on at least two of the following defined as: a 14% [0.6 SD] improvement in the physical function subscale of the SF-36; reduction in severity of the most troubling symptom endorsed by the patient to by at least 2 points on the visual analog scale; no longer a “fatigue case” on the Chalder; much improved or very much improved [+2 or +3] on the PGIC.

### *A priori* criterion for secondary outcome

Improvement by at least 10 points on the short version of the POMS.

### *A priori* criterion for successful outcome

At least a third of patients fulfill criteria for primary outcome.

## RESULTS

Two patients were lost to follow up leaving 14 for analysis. There were no adverse events reported during the course of this study. Table 1 shows the results.

**TABLE 1.** List of Criteria Successfully Attained by Subject.

| **SUBJ** | **PGIC** | **SF-36/PF Improved** | **Chalder Case-> No Case** | **VAS Improved** | **Positive Result** |
| --- | --- | --- | --- | --- | --- |
| 1 | 1 | 1 | 1 | 1 | 1 |
| 2 | 1 |  |  | 1 | 1 |
| 3 | 1 | 1 |  |  | 1 |
| 4 | 1 | 1 | 1 | 1 | 1 |
| 5 | 1 |  | 1 | 1 | 1 |
| 6 |  | 1 | 1 | 1 | 1 |
| 7 |  | 1 | 1 |  | 1 |
| 8 |  | 1 | 1 |  | 1 |
| 9 |  |  | 1 |  |  |
| 10 |  |  |  |  |  |
| 11 |  |  |  |  |  |
| 12 |  |  |  |  |  |
| 13 |  |  |  |  |  |
| 14 |  |  |  |  |  |

Two patients showed improvement on all 4 outcome measures; one on 3 outcome measures; and five on two of the outcome measures. Of the remaining patients, one improved on one outcome measure while 5 did not improve on any of the outcome variables. The outcome measures that best predicted a successful outcome were improvement on the Global Clinical Assessment of Change and/or improvement in at least one symptom on the visual analog scale. For patients who did not show improvement on these outcome measures, showing improvement on both the SF-36 Physical Function and loss of caseness on the Chalder was the next predictor of a successful outcome. Of the symptoms captured on the visual analog scales, brain fog was the one symptom that showed no improvement for any patient.

In summary, 8 of the 14 patients fulfilled our *a* priori criteria for improvement. This result exceeds our *a priori* criteria of at least a third of patients improving. Thus, this was a successful pilot trial. Four of the patients fulfilling criteria for successful improvement plus one who did not show improvement had reductions of at least 10 points on the POMS.

## DISCUSSION

Long Covid is very common and doctors have little other than supportive care to offer such patients. This open label pilot study suggests non-invasive stimulation of the auricular branch of the vagus nerve is a possible therapeutic modality for treating this ailment. Since it did not have a sham stimulated group for comparison, it is possible that the positive result was simply a placebo response to treatment. Against that interpretation is the fact that a review of treatment trials in ME/CFS noted the highest placebo rate to be 24%^10^ while this study showed a 57% improvement rate.

There are important differences between the design we followed here and that used in most drug trials for medically unexplained widespread pain, known as fibromyalgia. In order to test the efficacy of their drug, these trials often require subjects to be off any brain-active medication^11^. We imposed no such criterion in this trial except to ask patients not to increase current medication dosages or start new ones. Doing this expands the number of patients willing and eligible to be in a trial. Another difference is that drug trials often define a successful outcome as improvement in a single measure of pain. In our earlier study of implanted VNS as a possible treatment of FM^12^, we required subjects improve on more than one outcome measure – a tactic we employed here. One might view our including only Long Covid patients fulfilling the 1994 case definition for CFS as a limitation. But we see advantages in making this choice: it reduces patient pool heterogeneity and selects patients with a broad set of symptoms at substantial or greater severity including brain fog, fatigue, widespread pain and post-exertional malaise.

Our obvious next step is to expand this study and to add a sham control limb in order to assure the efficacy of VNS in treating Long Covid. In such a study we would no longer use brain fog as an outcome variable on a visual analog outcome variable. Should such a trial again show efficacy, this will open another avenue of treatment in the care of these patients for whom little specific treatment is currently available.

## Data Availability

All data produced in the present study are available upon reasonable request to the authors,

## Acknowledgement

This work was supported by donations from patients of Dr Natelson to support the activities of his laboratory. The devices were donated by the manufacturer, Parasym.

